# Knowledge, Attitude, and Practices of Health Professionals Working in A Major Health Care System Regarding Social Determinants of Health (SDOH) and Community Health Workers (CHW’s)

**DOI:** 10.1101/2022.02.21.22271311

**Authors:** Muhammad Nausherwan Khan, Erum Azhar, Areeba Zain, Meredith Root Bowman, Natasha Tahir Ahmed, Muhammad Arslan Khan, Leigh Caswell, Abdul Waheed

## Abstract

**Introduction:** Social Determinants of Health (SDOH) play a key role in impacting the health outcomes of any population. Community Health Workers (CHW’s) play an important role in health promotion, disease prevention, and management of chronic illnesses. This study aims at exploring the knowledge, attitude, and practices of health care professionals towards CHWs to fully integrate in them for mitigation of SDOH.

**Materials and Methods:** A cross-sectional study utilizing an anonymous survey questionnaire across 4 clinical sites was carried out from June 2016 to November 2017 in a major healthcare system (Presbyterian) in Albuquerque, New Mexico. Descriptive statistics (means, standard deviations, and proportions) were collected. Categorical variables were analyzed using Chi-squared and Fisher’s exact test; a p-value of <0.05 was considered statistically significant, using SAS 9.4 statistical software.

**Results:** Almost half of the health professionals had no knowledge about the social determinants of health. Almost a quarter of the health professionals did not know the role of CHWs in healthcare, however, 100% of the respondents across all clinic and practice locations and regardless of their role or scope of the practice believed that greater involvement of CHWs would improve patient outcomes.

**Conclusion:** There is a knowledge deficit among health care providers about the social determinants of health (SDOH).More educational and teaching opportunities on SDOH and CHWs to all health professionals should be provided to all health professionals so the clinical team can help manage SDOH in addition to providing clinical care.

## Introduction

Clinical care including access and quality of health care accounts for twenty percent of the health outcomes for a given population. The remaining 80% is impacted by health behaviors, social and economic factors as well as physical environments [1]. Someone must take responsibility to address the remaining 80% of health factors that impact the health outcomes of a population. Literature suggests that nearly half of the primary care physicians have already reported burnout rates [2]. It is pertinent to broaden the health care team to assist with the management of social determinants of health (SDOH) which ultimately impact patients’ health outcomes. Currently, there is a supportive health care climate for primary care physicians, allowing for investment and funding. Adding the community care workers to the clinical care team or the PCMH (Patient-Centered Medical Home) and managing it under the primary care provider, is an important step that can impact the social determinants of health (SDOH) effectively [3].

There was a time when diagnosing and treating patients’ disease processes was considered adequate when it came to primary care clinics. However, while dealing with patient care, primary care physicians deal with numerous added responsibilities in a health care system including and not limited to offering preventive services, controlling chronic conditions, providing mental health to bridge the gap of limited mental health care providers, coordinating with numerous community and law agencies, filling numerous forms and documentation on the electronic medical records as well as leading and organizing multidisciplinary teams for patient care. On the other hand, social determinants of health itself are frequent and problematic, needing extra time and adding to the stress along with providing traditional medical services leading to higher burnout rates among providers [4].

Though the majority of the physicians believe that SDOH matters to their patients, they do not believe it is their direct responsibility to address the SDOH with the patients. Others do not quite understand how to tackle complex issues related to social determinants. Thus, strategies to address these social needs of the patients must first acknowledge the existing barriers that limit physician’s ability to help patients with these needs, at the same time avoiding physicians’ burnout and overburdening them [5]. Some strategies suggested for overcoming these barriers are to promote an effort to help clinical sites address patients’ SDOH without contributing to physician burnout. Engaging employer and policymaker as a key stakeholder in efforts to improve community health. Increasing investment in public health would also help resolve these barriers [5]. Three Family Medicine clinics in Albuquerque, New Mexico, staffed by Family Medicine attendings and four residents, screened 3,048 patients for SDOH and found that nearly half (46%) screened positive for at least one social need. Community health workers would then help patients with appropriate community resources [6]. One randomized control trial of CHWs supporting low-income patients with multiple chronic conditions demonstrated improvements in health outcomes, mental health, and reductions in hospitalizations [7]. Similarly, a systematic review revealed that CHW interventions can significantly reduce emergency room visits, hospitalizations, and urgent care visits among patients served in various healthcare settings[8]. Overall, there are known positive effects of community health workers (CHW’s) on addressing social determinants of health, improving patient health outcomes, and decreasing overall healthcare costs. To our knowledge, there is limited literature available, and not many studies have been done to study the knowledge, attitudes, and practices of health professionals towards SDOH and CHWs. This study aims at exploring the knowledge, attitude, and practices of health care professionals towards CHWs which in our opinion is the first step towards CHW’s greater involvement in clinical practice and addressing SDOH.

## Materials and Methods

After approval by the institutional review board (IRB), all health care professionals who had an impact on SDOH were included in the study population. This included health care providers physicians, advanced practice clinicians (APCs), nurses, medical assistants (MA’s), social workers, care coordinators, and other ancillary staff. Four clinical sites were included in the study: PMG Isleta clinic, Kaseman IM clinic, Kaseman ED, and Kaseman IP Psychiatry.

It was a cross-sectional study utilizing an anonymous survey questionnaire disseminated from November 2017 to June 2016. Convenience sampling was used. The survey consisted of 15 questions. The questionnaire elicited information regarding demographics (4 questions), knowledge about the social detriment of health and community health workers (3 questions), and social issues reported by patients in the practices (5 questions), and resources to refer to and any outcome that is affected by utilizing CHW. The multiple-choice response format was used for most questions, with options of never, almost never, occasionally /sometimes, almost every time, and every time.

Data was entered on Microsoft Excel. Descriptive statistics (means, standard deviations, and proportions) were collected. Categorical variables were analyzed using Chi-squared and Fisher’s exact test; a p-value of <0.05 was considered statistically significant, using SAS 9.4 statistical software.

## Results

Survey was done at four different locations and a total of one hundred and twenty two medical staff members responded. Table one shows socio-demographic characteristics of the respondents. The sites where the majority of the health professionals respondents filled our survey namely Kaseman ED (ED) and Kaseman inpatient and Behavioral Health (BH) reported a heavy burden of low-income patients with social determinants of health (SDOH) in their patient encounter in these practice sites/locations with Kaseman ED having an encounter of Every time/ almost every time 92.31% (n=48), whereas, Kaseman inpatient and outpatient Behavioral health reported having encounter of every time/almost every time as 83.33% (n=25) (Figure 1).

**Figure 1:**
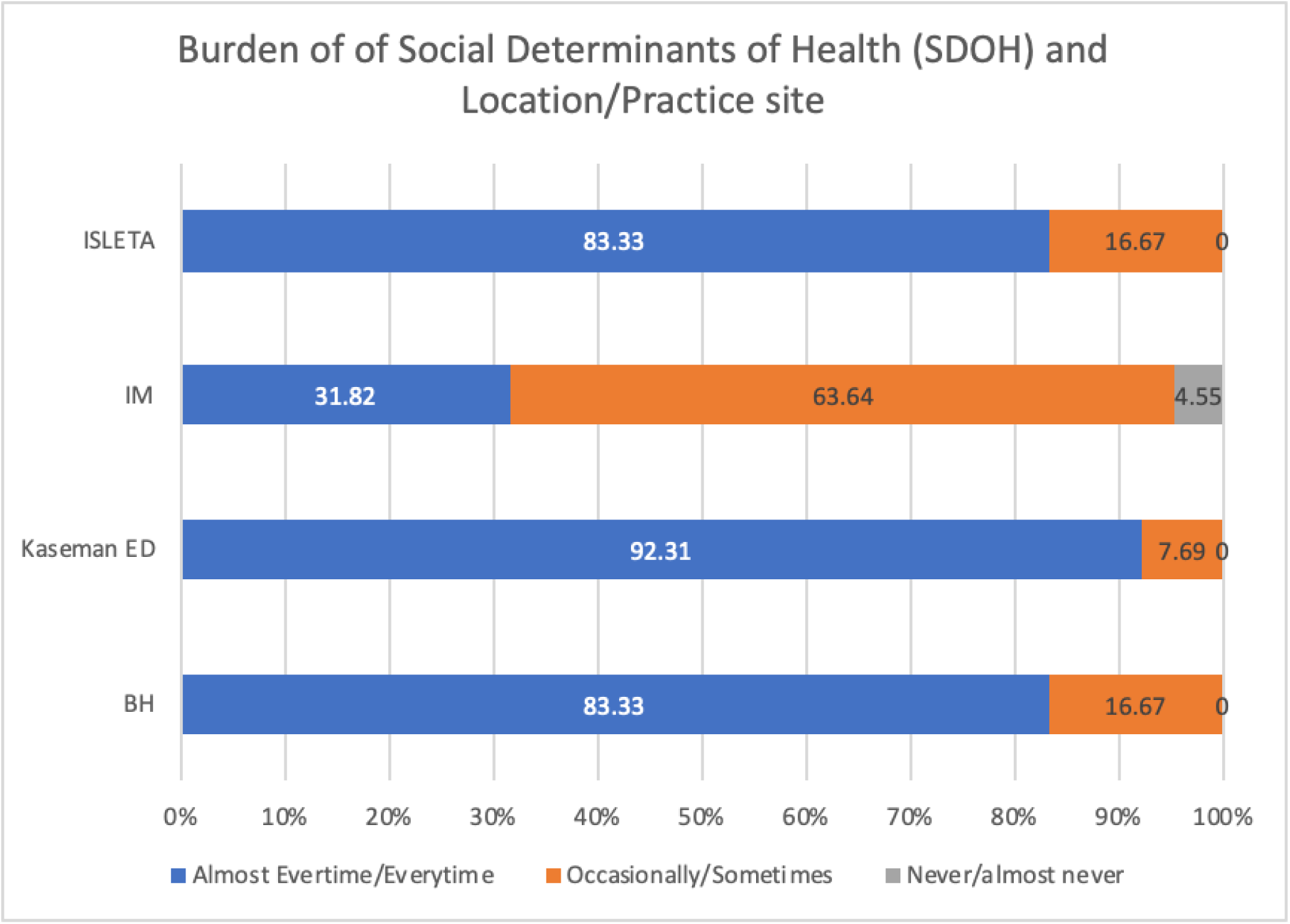
Burden of Low-Income Patients and Social Determinants of Health (SDOH) at Different Practice Site.

**Table 1:** The Socio-Demographic Characteristics Of The Survey Respondents.

Respondents
| Gender (missing n=1) | N (total n=122) | % |
| --- | --- | --- |
| Male | 39 | 32.23% |
| Female | 82 | 67.77% |
| Age (missing n=5) | N | % |
| 20-29 | 15 | 12.82 |
| 30-39 | 35 | 29.92 |
| 40-49 | 32 | 27.35 |
| 50-59 | 15 | 12.82 |
| 60-69 | 19 | 16.24 |
| 70-79 | 1 | 0.86 |
| **Practice Area** | **N** | **%** |
| Physician | 26 | 21.31 |
| APC (PA or NP) | 20 | 16.39 |
| Nurse (RN) | 29 | 23.77 |
| MA | 9 | 7.38 |
| Clinic Manager or Administrator | 3 | 2.46 |
| Care Coordinator or Social Worker | 4 | 3.28 |
| Others | 31 | 25.41 |
| **Years of Practice (missing n=6)** | **N** | **%** |
| Less than 2 Years | 2 | 1.72 |
| 2-4.9 Years | 24 | 20.69 |
| 5-9.9 Years | 14 | 12.07 |
| 10-19.9 Years | 36 | 31.03 |
| 20-29.9 Years | 22 | 18.97 |
| Over 30 Years | 18 | 15.52 |
| **Practice Locations** | **N** | **%** |
| Kaseman inpatient and outpatient<br>Behavioral Health (BH) | 30 | 24.59 |
| Kaseman ED (ED) | 52 | 42.62 |
| Kaseman Internal Medicine PMG (IM) | 22 | 18.03 |
| Isleta PMG (Isleta) | 18 | 14.75 |

Almost half of the responding medical staff members 44.26% (n=54) did not know about social determinants of health, however, of those who did not know about the SDOH, 76.92% (n=50) showed interest to get further information about the SDOH. Among the survey respondents, 83.33% (n=100) knew who to contact if any social issues are identified in a patient. as shown in Figure 2.

**Figure 2:**
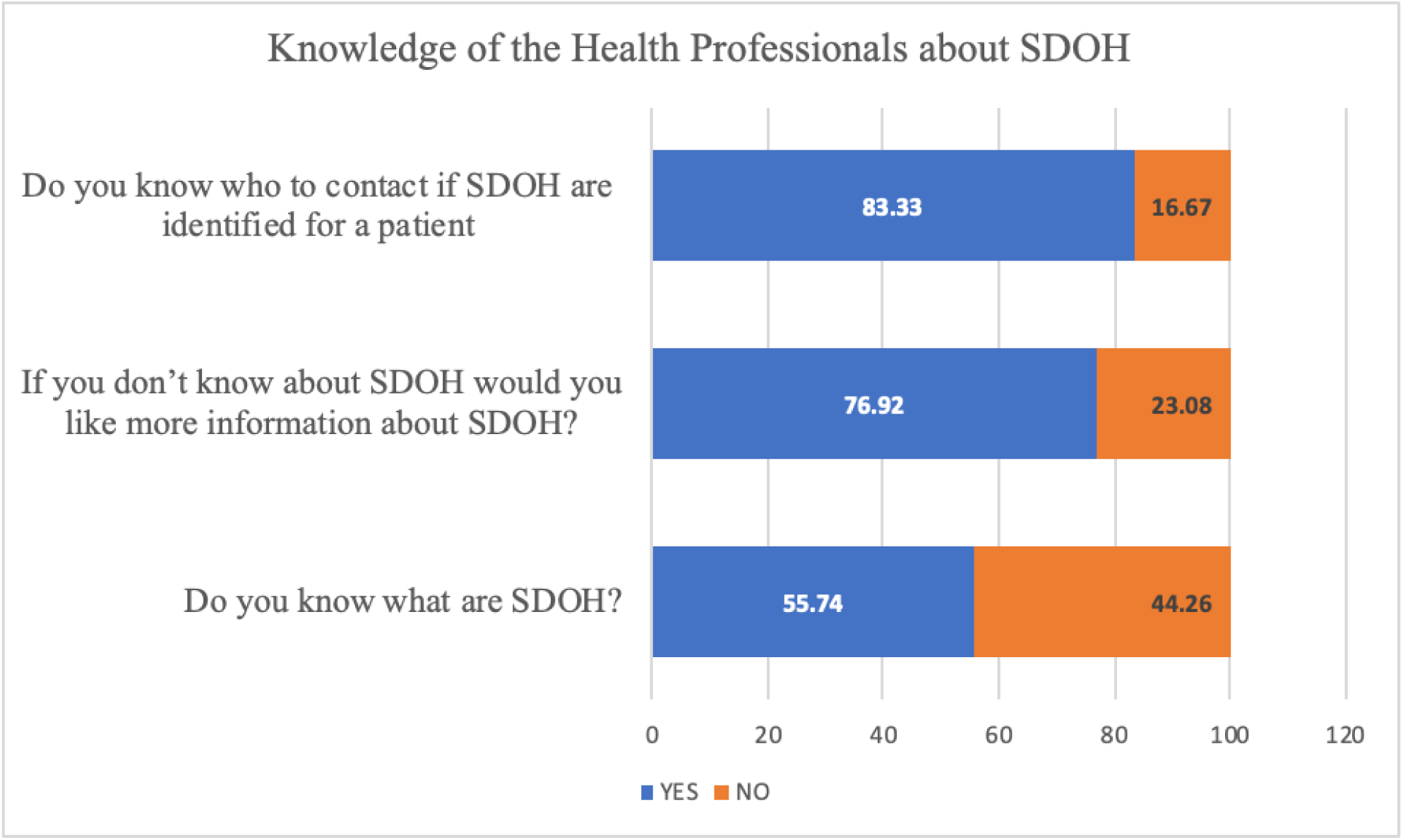
Knowledge of Health Professionals about Social Determinants of Health (SDOH)

The knowledge about SDOH was not different among medical staff members (Physicians, Advanced practice manager (APC) that included physician assistant (PA) and Nurse practitioner (NP), clinic manager/administrator, behavioral health practitioner, clerical and Emergency room technician) (p=0.1406). Of the 44.26% (n=54) medical staff members who were not aware of the social determinants of health, the attitude to learn about it was mostly positive and 76.92% (n=50) would opt to have further information about social determinants of health. This positive attitude was seen across all members of the medical staff and there was no difference between medical staff members and scope of practice (Physicians, APC(NP+PA), Nurses (RN), and Allied health/others (p=0.58). Hundred Medical staff members (83.33%) knew who to contact if a patient with social determinants of health is encountered. This knowledge was not statistically varied among different medical staff members (p=0.66). Among them, 38% (n=38) were health providers (Physicians+APC’s) and 62% (n=62) were other medical staff members, and further analysis did not reveal any difference between providers (physicians +APC) and other combined medical staff members (p= 0.80). The majority would contact a case manager or case coordinator 72.13% (n=88) of the times, social workers were contacted 68% of the times (n=83) and community health workers would be contacted only 18.85% of the times(n=23). Among the survey respondents, 83.33% (n=100) knew who to contact if any social issues are identified in a patient, 45% were from ED location, followed by BH 22%, 18% IM, and 15% Isleta, however, this difference in knowledge was not statistically different at various locations (p=0.64).

Of the 55.74% (n=68) of the medical respondents who reported being knowledgeable about the SDOH, the ED site reported being the most knowledgeable about SDOH with 38.24% (n=26), followed by the BH site’s knowledge 25% (n=17) not shown in figure, however, there was no statistically significant association seen regarding knowledge about social determinants of health (SDOH) and practice site/location (p= 0.23) as shown in Figure 3.

**Figure 3:**
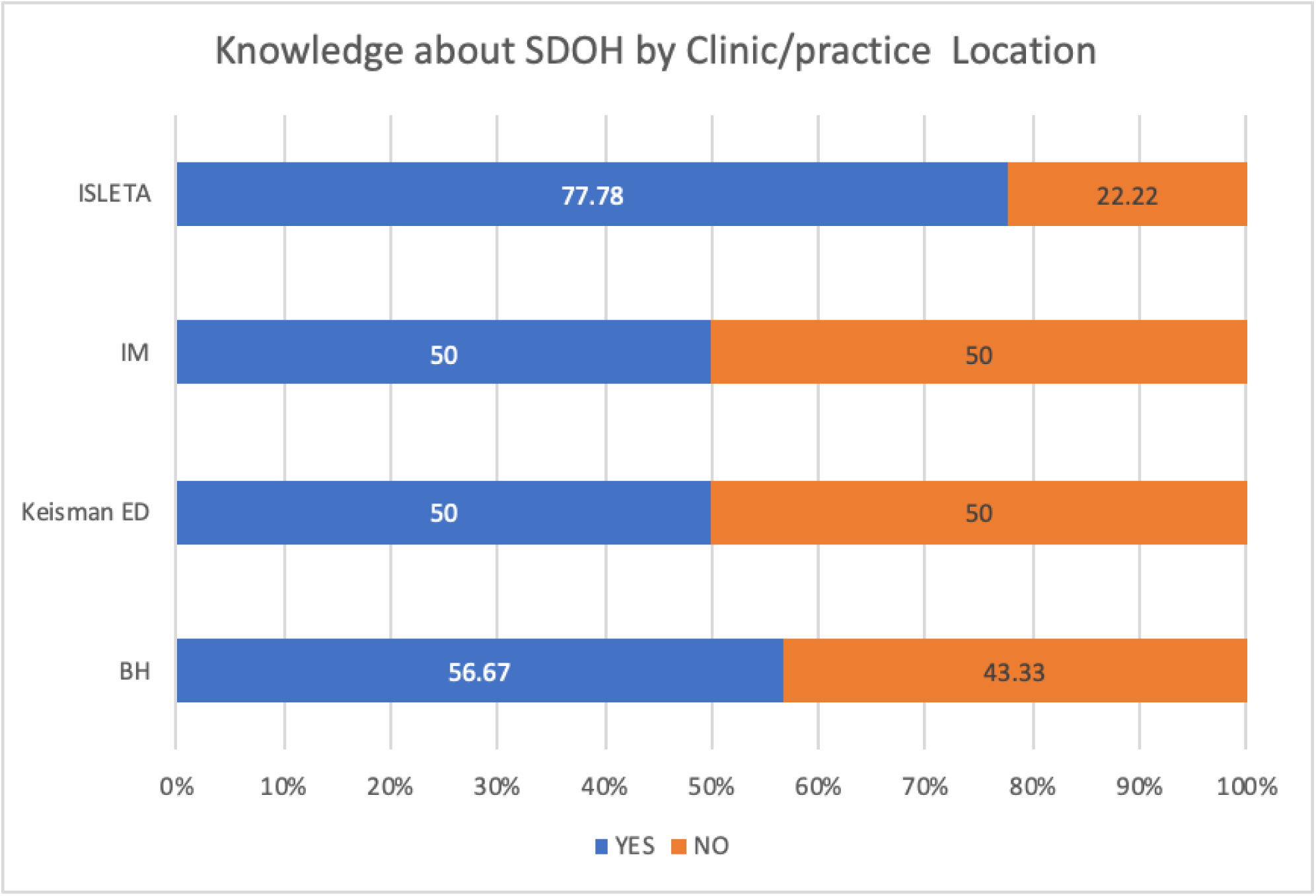
Knowledge about Social Determinants of Health by Clinic/Practice location.

A significant number of medical staff members encounter patients with SDOH issues “occasionally /sometimes” by 21.31% (n=26) to “almost every time” by 59.02% (n=72). Housing and other utilities issues were mentioned by patients to these medical staff members “occasionally /sometimes “by 69.67% (n= 85) to “almost every time” by 13.93% (n=17).

Medical staff members were approached by patients regarding food insecurities “occasionally/sometimes” by 59.84% (n=7). Patients mentioned transportation issues to these medical staff members “occasionally/ sometimes” by 68.85% (n=84) to “almost every time” by 24.59%(n=30) patients. Physical or emotional abuse (violence) at home was reported to these responding medical staff members “occasionally/ sometimes” by 68.85% (n=84) of the patients as shown in Figure 4.

**Figure 4:**
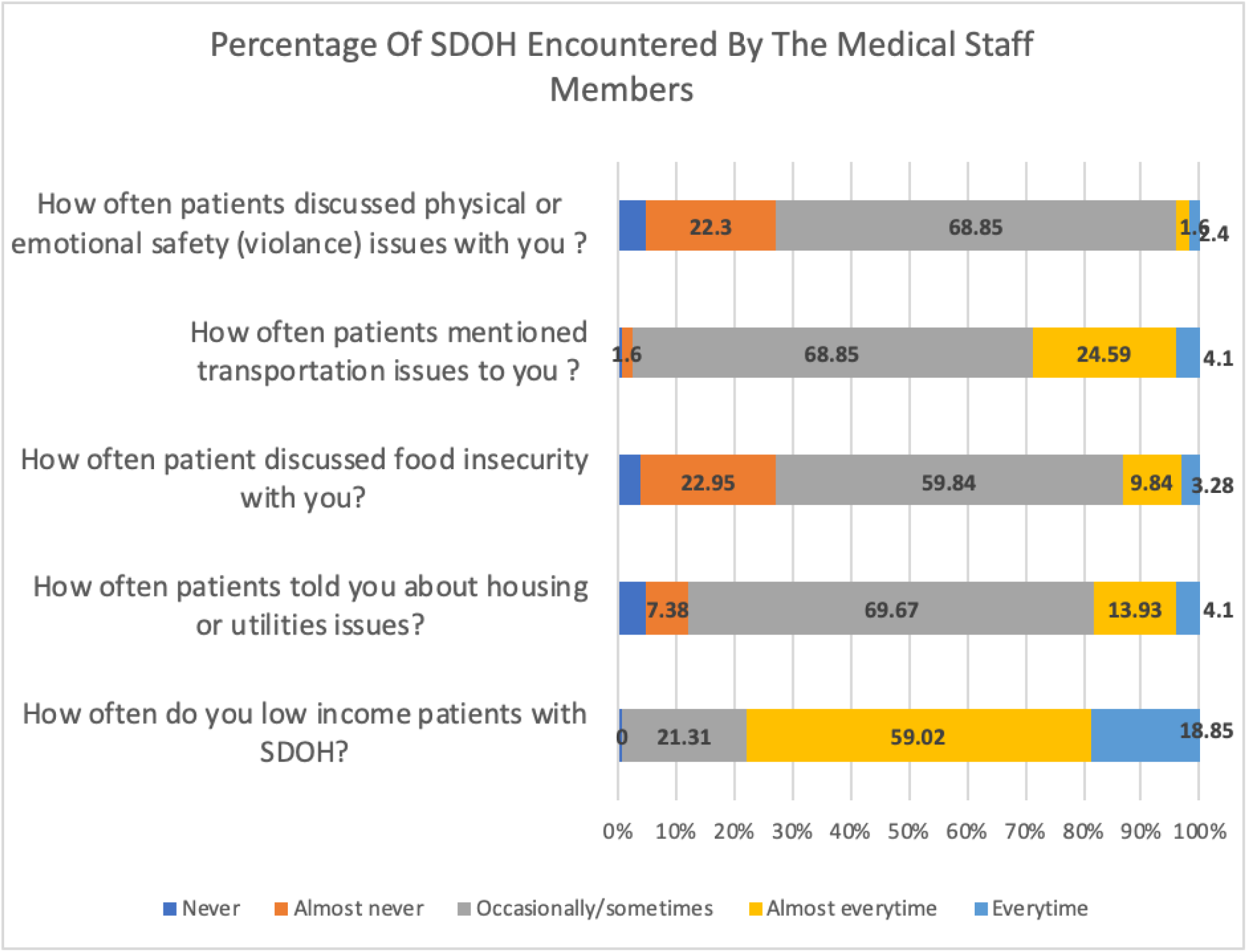
Frequency of SDOH Medical Staff Members Encounter of SDOH, Utilities, Food Insecurity, Transportation, and Violence.

Social determinants of health SDOH responses encountered by the responding medical staff members were not statistically different between males and females’ gender of the members of the medical staff (p=0.89). However, we noted more patient encounters reporting transportation issues to more female staff members as compared to male medical staff members with a p-value of 0.0135.

There was no difference in responses for the SDOH between those medical staff members who have been in practice for five or more years versus those who have been in practice for less than five years (p= 0.69). Encounters reported by patients regarding housing/utility issues (p=0.17), food insecurity (p=0.27), transportation issues (p=0.165) and physical and emotional safety (violence)issues (p=0.08) were also not statistically different among medical staff members who have been in practice for five or more years versus those who have been in practice for less than five years.

Social determinants of health SDOH encountered by the responding medical staff members were not statistically different among different members of the medical staff and their scope of practices (Physicians, Advanced practice manager (APC) that included physician assistant (PA) and Nurse practitioner (NP), Allied health/others (including medical assistant (MA), social worker/care coordinator and others including clinic manager/administrator, behavioral health practitioner, clerical and Emergency room technician) (p=0.84). Encounters reported by patients regarding housing/utility issues (p=0.73), food insecurity (p=0.17), transportation issues (p=0.21) and physical and emotional safety (violence)issues (p=0.94) were also not statistically different among different health professionals and their scope of practices as shown in Figure 5.

**Figure 5:**
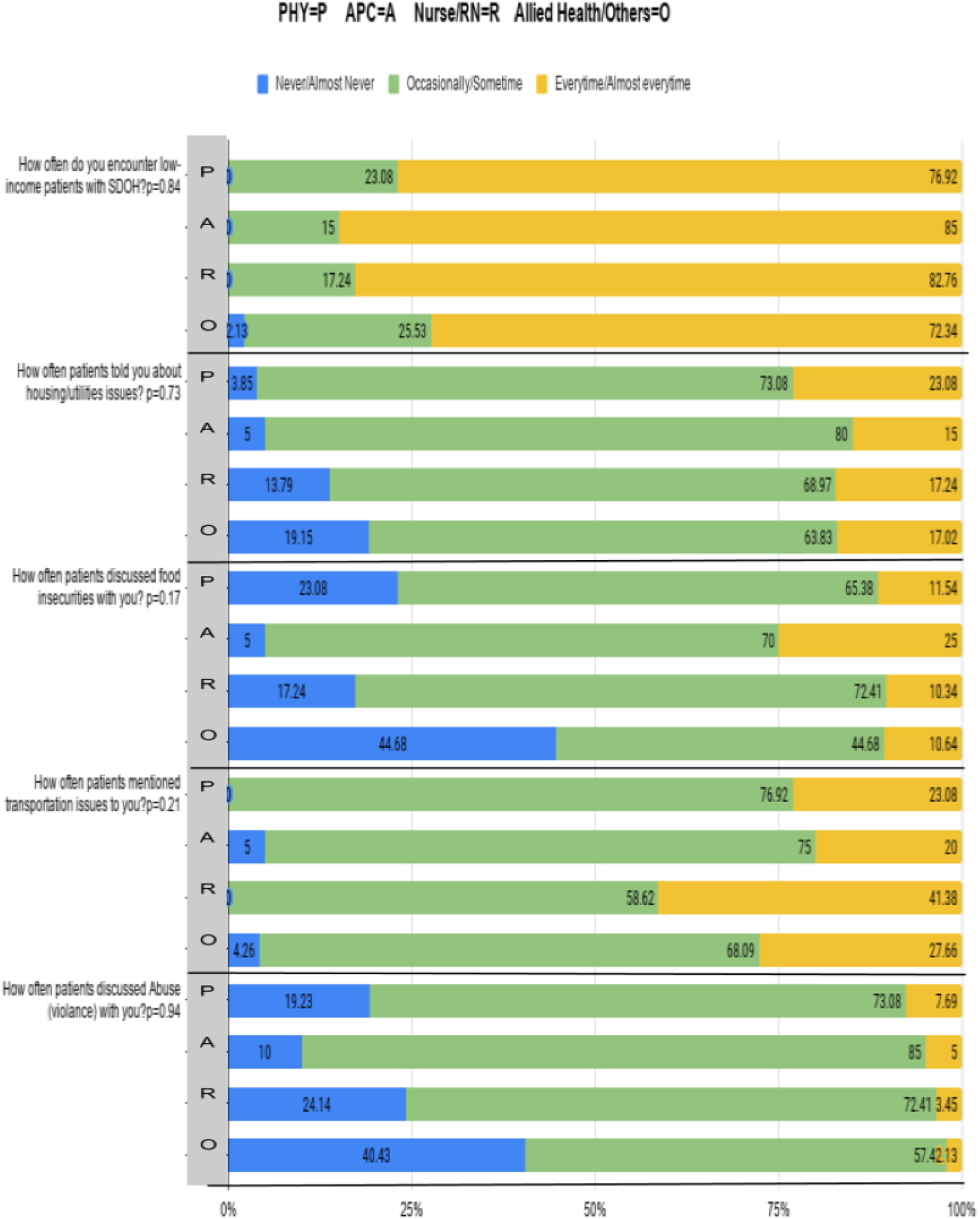
Scope of Practice and SDOH Reported by Patients to the Medical Staff Members.

Low-income patients with SDOH were encountered every time/almost every time at a greater percentage at the location sites of Kaseman ED (ED) (92.31%), Kaseman inpatient and outpatient Behavioral Health (BH) (83.33%), and Isleta (83.33%) compared to IM practice site (31.82%) with statistically significant p-value 0.03. There was no significant association seen between different practice sites and encounters by the medical staff for patients with utilities/housing issues (p=0.51), food insecurity (p=0.24), or transportation issues (p=0.899), however, significantly more medical staff encountered “occasionally /sometimes” patients who reported domestic violence at BH site (86.67%) and ED site (82.69%) compared to the other practice sites of IM (36.36%) and Isleta (38.89%) with a significant p-value of 0.0001 as shown in Figure 6.

**Figure 6:**
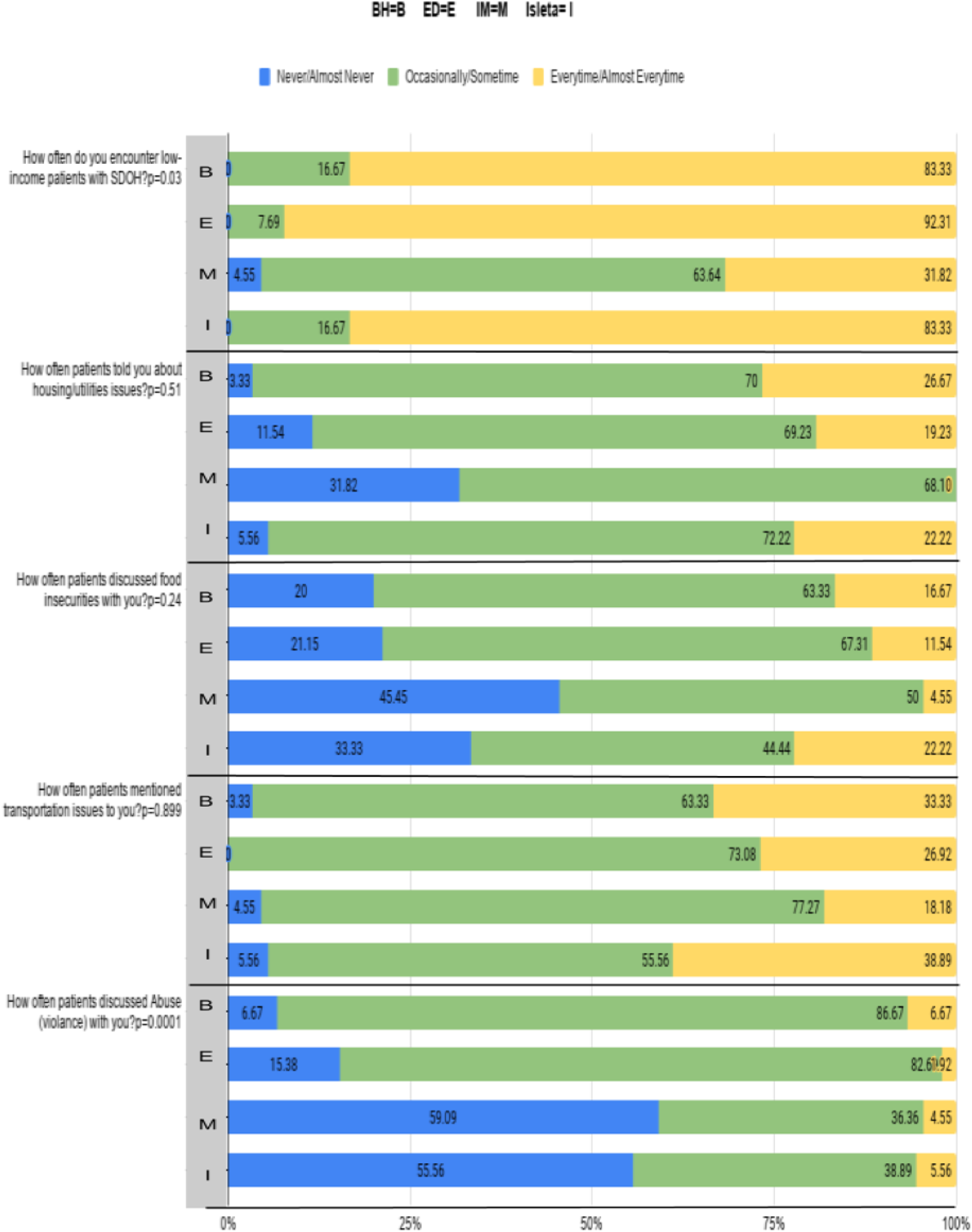
Practice Site and SDOH Reported by the Patients to the Medical Staff Members.

It was interesting to find that 73.33% (n=87) of the medical staff members that responded had never worked with a CHW before and 22.13% (n=27) of the respondents were not even aware of what a CHW does. However, 89.08% (n=106) showed interest to know more about Community Health Workers (CHW’s) as shown in Figure 7.

**Figure 7:**
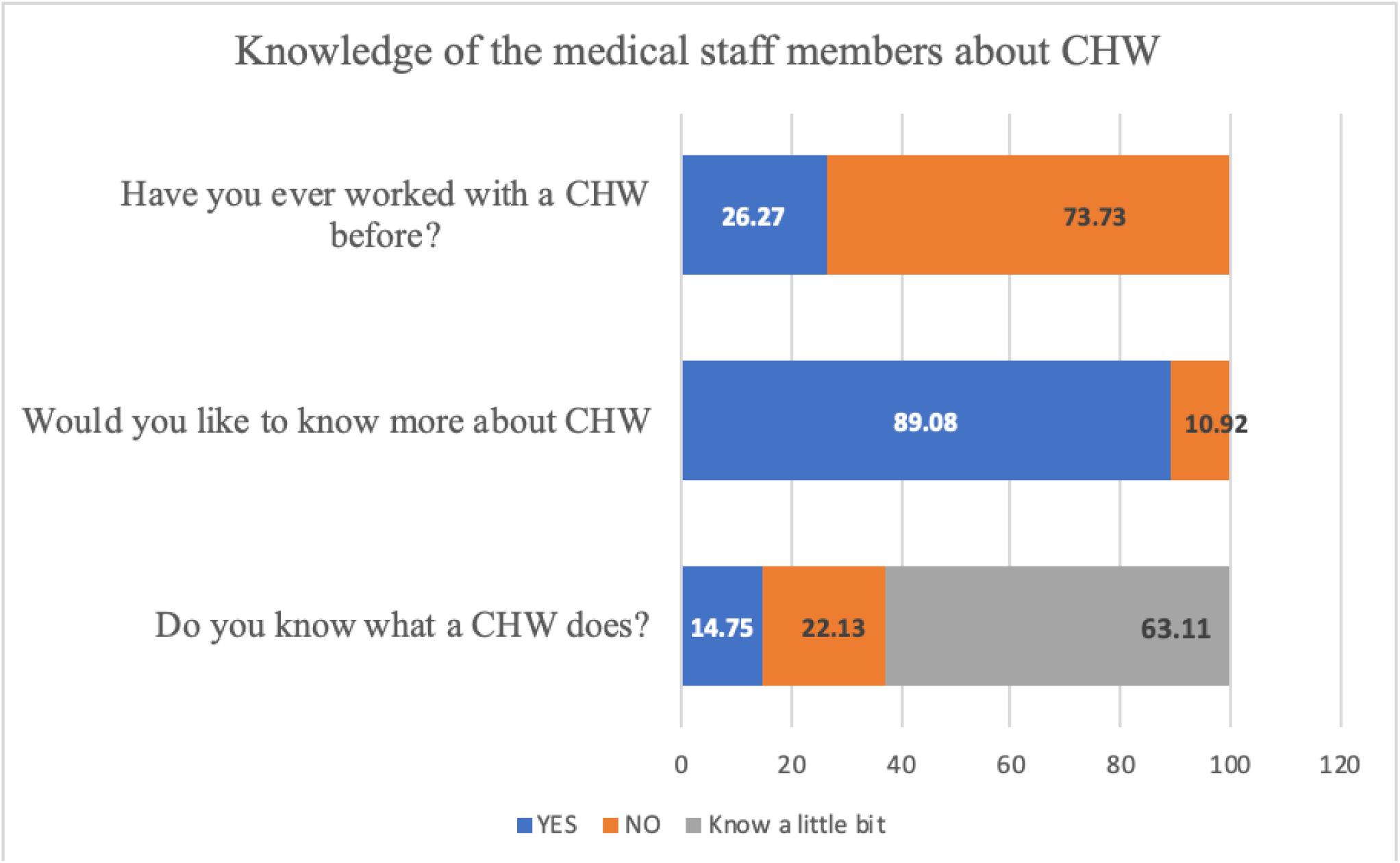
Knowledge of Medical Staff Members about Community Health Workers (CHW).

There was also no correlation between the scope of practice (physician, APC, RN, Allied health/others) and their knowledge of the role of community health workers (CHW’s) with a p-value of 0.40. Of the 10.92% (n=13) medical staff members who were not interested in knowing more about CHW, 7.69% were health providers and 92.31% were other medical staff members, thus more other staff members were not interested in knowing more about CHW as compared to health providers with a significant p-value of 0.0176. Of the 73.33% of the respondents who have not worked with the community health workers, 40.23%% were health providers and 59.77% were other staff members with no statistical difference seen between different providers and working with CHW (p=0.15). Of the total 26.27% respondents who have worked with the community health workers before, the majority were from the Isleta location (45.16%), followed by BH 22.58% and the remaining 16.13% each from ED and IM location, and this difference was statistically significant, so more medical staff members at Isleta and BH have worked with CHW as compared to ED and IM location(p-value 0.0001).

A hundred percent of the responding medical staff members thought that patient outcomes would improve if a CHW was involved in the care of the patient regardless of their scope of practice or providers versus non-providers. There was also no difference between practice location/site and all staff members at all locations agreed to a hundred percent that utilizing community health workers (CHW’s) services in the care of the patient would improve patient outcomes.

## Discussion

There is a knowledge deficit among health care providers about the social determinants of health (SDOH). Almost half of the responding medical staff catering to the population with heavy SDOH were not familiar with the social determinants of health. This is a significant number of healthcare workers which shows that our health systems are ill-equipped to deal with SDOH. This knowledge deficit is also shown by similar studies. A study published in Canada in 2016 reported that providers often felt helpless and frustrated when faced with the complex and intertwined health and social challenges of their patients and as a result many avoid asking about social issues, preferring to focus on medical treatment and lifestyle counseling [9]. Considering this, there has been a call for greater emphasis on the social accountability of medical schools and other institutions responsible for training health professionals to better cater to disadvantaged patients. Certain changes can be considered for implementation including tests for knowledge of social determinants of health in licensure exams, continuing education courses, and in other credentialing capacities for health care workers. Despite having limited baseline knowledge about SDOH, our study found that the participants showed a willingness to learn more. This is consistent with previously published literature[10]. Girgis et al report that despite limited resources, physicians were willing to help their patients through social challenges [10]. Our study shows that of the approximately 10% of respondents who reported as not interested in knowing more about CHWs, of them 92.3% identified themselves as other medical staff and 7.7% were health providers. This is an encouraging finding as personnel responsible for providing direct patient care and with the responsibility of initiating interventions form a small minority of people who responded not be interested to learn more regarding the role of CHWs.

It was also seen that the knowledge about SDOH was not dependent upon the role such as between providers and other medical staff. Our study also demonstrated that the burden of social determinants of health was present most of the time in varying degrees across the four practice locations. It was also interesting to see that ED was the most knowledgeable when it came to knowledge about SDOH. Perhaps this is no surprise as ED providers are responsible for caring for a large volume of vulnerable patients who seek refuge EDs around the country [11]. In addition to this, it was intriguing to see that patients disclosed issues with transport more to female health care staff as compared to males. There was no difference in the number of years of practice or dependent on the level/scope of practice. This means a level ground is present and education and awareness efforts can be directed uniformly towards all providers regardless of years of experience or specialty.

Our study also found that once healthcare workers identified patients with social issues, 83.3% thought they knew who to contact to address the patient’s concerns. In the majority of cases, this person was either the case manager or case coordinator (72.13%) or social workers (68%). CHWs were only contacted in a minority (18.85%) of instances. This finding can be related to the fact that the majority of the staff (73.3%) that had responded had never worked with the CHW before and a minority (22%) did not even know what a CHW did. This is a very important finding from our study and shows the acute need for healthcare workers to be educated in regards to the potential role CHWs can play in the improvement of overall health outcomes of our patients.

In 2013, the Affordable Care Act (ACA) recognized the Community Health Workers (CHWs) as frontline public health workers and distinct members of the health care team. Subsequently in 2014, the Centers for Medicaid and Medicare (CMS) put forth guidelines for reimbursement of preventive services offered by CHWs. [12] These two developments laid the groundwork for greater integration of CHWs into the primary care structure. In addition to this, a greater role for CHWs in primary care setting helps achieve the Institute for Healthcare Triple Aim to improve the patient experience of care (including quality and satisfaction), improve the health of populations, and reduce the per capita cost of health [13]. A multicenter randomized clinical trial of 592 adults, studied the effect of Community Health Worker support on clinical outcomes of low-income patients across primary care facilities and found that the participants reported greater quality of primary care when CHWs were involved in the care[14]. Another study found that support from CHWs (vs goal-setting alone) led to improvements in several chronic diseases [7]. CDC has outlined the role of CHWs in chronic disease prevention and health promotion as well [15,16]. With half of the responding medical staff catering to the population with heavy SDOH not familiar with the social determinants of health, our study sheds light at a major barrier for integration of CHWs into the primary care landscape. There is a need to familiarize the healthcare workforce with importance of CHWs and their role and to set systems in place to follow guidance from CDC and CMS for this integration to take place. Another important finding from our study shows that all respondents of the study unanimously agreed regarding the benefit of involving CHWs in the care to improve patient outcomes.. The fact that a hundred percent of the respondents across all locations and regardless of their role or scope of the practice believe that greater involvement of CHWs would only benefit patients is perhaps the first step towards the expansion of the role CHWs play in patient health outcomes in the future. Our study has both strengths and limitations. This study has been able to identify current knowledge, attitudes, and practices for the Presbyterian healthcare system regarding SDOH and CHWs. One of the strengths of the study is that it was carried out at multiple sites catering to a wide population and included physicians, APCs, and Allied health workers. Limitations include impact by respondents underlying contextual and cultural factors that cannot be accounted for accurately.

Further studies could focus on improvement in knowledge, attitudes, and practices. The study provided an opportunity to not only gather data but to introduce the Accountable Health Communities CMS project, the center for community health and population health, and to answer any related questions, thus providing an opportunity for education and teaching. Our survey and findings of the study can be utilized for future quality improvement studies to improve the integration of CHWs in the primary care setting.

## Conclusion

Our study findings are a cause for concern because of knowledge deficit in the health care professionals regarding CHWs. More educational and teaching opportunities on SDOH and CHWs to all health professionals should be provided, including CME’s and posters/flyers. Education is important for all health professionals, not just health providers, since the whole clinical team is involved in maintaining the functionality of a PCMH and ensuring an environment that can help manage SDOH in addition to providing clinical care. Future studies are needed to accurately identify gaps in knowledge. Post-intervention studies would be of benefit to find out the impact of teaching and educational opportunities.

## Data Availability

All relevant data are within the manuscript and its Supporting Information files.

## Acknowledgements

We would like to acknowledge Dr. Darcie Robran-Marquez (Medical Director of Population Health at Presbyterian Healthcare System), Dr. Jason Mitchell (CMO of Presbyterian Healthcare System), Dr. Fernando Jumalon (Medical Director of Presbyterian Hospital Adult Internal Medicine Service), and the entire Community Health Department at Presbyterian Healthcare System, who fully supported our team during this research study.

## Notes

### Competing Interest Statement

The authors have declared no competing interest.

### Funding Statement

The author(s) received no specific funding for this work.

### Author Declarations

All experimental protocols were approved by Presbyterian Hospital and health system IRB, Albuquerque NM.

## References

1. County Health Rankings Model (2018)Collaboration Between University of Wisconsin Population Health Institute And Robert Wood Johnson Foundation.Http://www.Countyhealthrankings.Org/County-Health-Rankingsmodel

2. West CP, Dyrbye LN, Shanafelt TD. Physician burnout: contributors, consequences, and solutions. J Intern Med. 2018 Jun;283(6):516–529. DOI: 10.1111/joim.12752. Epub 2018 Mar 24. PMID: 29505159.

3. Kaufman A. Theory vs Practice: Should Primary Care Practice Take on Social Determinants of Health Now? Yes. Ann Fam Med. 2016 Mar;14(2):100–1. DOI: 10.1370/afm.1915. PMID: 26951582; PMCID: PMC4781510.

4. Solberg LI. Theory vs Practice: Should Primary Care Practice Take on Social Determinants of Health Now? No. Ann Fam Med. 2016;14(2):102-103. DOI:10.1370/afm.1918

5. Winfield L, Desalvo K, Muhlestein D. Leavitt Partners. Social Determinants Matter, But Who Is Responsible? 2017 Physician Survey on Social Determinants Of Health.

6. Page-Reeves J, Kaufman W, Bleecker M, Norris J, Mccalmont K, Ianakieva V, etal. Addressing Social Determinants of Health In A Clinic Setting: The Wellrx Pilot In Albuquerque, New Mexico. J Am Board Fam Med. 2016 May 1;29(3):414–8.

7. Kangovi S, Mitra N, Grande D, Huo H, Smith R, Long J. Community health worker support for disadvantaged patients with multiple chronic diseases: a randomized clinical trial. Am J Public Health. 2017;107:1660–1667. doi: 10.2105/AJPH.2017.303985

8. Jack HE, Arabadjis SD, Sun L, Sullivan EE, Phillips RS. Impact of community health workers on use of healthcare services in the United States: a systematic review. J Gen Intern Med. 2016;32:325–344. doi: 10.1007/s11606-016-3922-9

9. Andermann A: Taking action on the social determinants of health in clinical practice: a framework for health professionals. Canadian Medical Association Journal. 2016, 188:E474– 83. 10.1503/cmaj.160177

10. Girgis L, Van Gurp G, Zakus D, et al.: Physician experiences and barriers to addressing the social determinants of health in the Eastern Mediterranean Region: a qualitative research study. BMC Health Services Research. 2018, 18:. 10.1186/s12913-018-3408-z

11. Anderson E, Lippert S, Newberry J, et al.: Addressing Social Determinants of Health from the Emergency Department through Social Emergency Medicine. Western Journal of Emergency Medicine. 2016, 17:487–9. 10.5811/westjem.2016.5.30240

12. IMPACT OF COMMUNITY HEALTH WORKERS IN PRIMARY CARE Arizona Health Care Provider Survey Results. 2015.

13. Balcazar H, Lee Rosenthal E, Nell Brownstein J, et al.: Community Health Workers Can Be a Public Health Force for Change in the United States: Three Actions for a New Paradigm. American Journal of Public Health. 2011, 101:2199–203. 10.2105/ajph.2011.300386

14. Kangovi S, Mitra N, Norton L, et al.: Effect of Community Health Worker Support on Clinical Outcomes of Low-Income Patients Across Primary Care Facilities. JAMA Internal Medicine. 2018, 178:1635. 10.1001/jamainternmed.2018.4630

15. CDC: Addressing Chronic Disease through Community Health Workers: A Policy and Systems-Level Approach. 2015.

16. How the Centers for Disease Control and Prevention (CDC) Supports Community Health Workers in Chronic Disease Prevention and Health Promotion. https://www.cdc.gov/dhdsp/programs/spha/docs/chw_summary.pdf

